# Prevalence and Determinants of Premarital Sexual Practice Among Youths in Ethiopia: Further analysis of the Ethiopian Demographic and Health Survey

**DOI:** 10.1101/2020.10.08.20209577

**Authors:** Kegnie Shitu Getie, Ayenew Kassie, Maereg Woldie

**Affiliations:** Department of Health Education and Behavioral Sciences, Institute of Public Health, University of Gondar, Gondar, Ethiopia

**Keywords:** Premarital Sex, Youths, Ethiopia

## Abstract

**Introduction:** Premarital sexual practice becomes a common phenomenon among youths in Ethiopia. It is usually associated with an unwanted pregnancy, abortion, and sexually transmitted diseases including HIV/AIDS.

**Objective:** This study aimed to assess the prevalence and determinants of premarital sexual practice among Ethiopian youths.

**Methods and Materials:** A Community-based cross-sectional study was conducted in all regions of Ethiopia from January 18 to June 27, 2016. All participants from the age of 15-24 (7, 389) were included for the analysis of the present study. Bivariable and multivariable binary logistic regression analytical models were fitted to identify factors associated with the premarital sexual practice. A 95% CI and p-value < 0.05 were used to declare statistical significance.

**Result:** The prevalence of premarital sexual practice was 10.8% (95% CI, 10 % – 11.5%). Being in the age group of 20 - 24 (AOR = 3.6, 95% CI (2.8, 4.6)), male sex (AOR = 1.7, 95% CI (1.3, 2.2)), employed (AOR = 1.4, 95% CI (1.03, 1.8)), from pastorals region (AOR= 1.4, 95% CI (1.3,2.4)), having mobile phone (AOR=1.7, 95% CI, (1.3, 2.3)), ever use of internet (AOR = 1.8, 95% CI (1.3, 2.5)), ever drinking alcohol AOR = 2.4, 95% CI (1.7, 2.5)), ever chewed khat (AOR = 2.4, 95% CI (1.6, 3.5), and ever tested for HIV (AOR = 1.3, 95% CI (1.1,1.6)) were a statistically significant factors associated with premarital sexual practice at p value less than 0.05.

**Conclusion:** For every 10 youths at least one of them had sexual intercourse before they got married. Being in the age group of 20 - 24, male sex, employed, from a pastoral region, having a mobile phone, ever use of the internet, alcohol drinking, khat chewing, and ever tested for HIV were significant factors associated with the premarital sexual practice. Thus, national sexual education and reproductive health behavior change interventions should give due attention for those groups. Indeed, adequate education should be given about premarital sexual intercourse when youths come for HIV test.

## Introduction

In 2019, about 1.2 billion people in the world were between the ages of 15 and 24. More than 200 million live in sub-Saharan Africa, where the largest increase in the proportion of youths is expected in the coming three decades [1]. In Ethiopia, youths aged 15–24 years are more than 15.2 million, contributing to 20.6 % of the whole population [2].

This large and productive group of the population is in a state of rapid physical and psychological changes with curiosity and the urge to experience new phenomena including sexual intercourse [3]. These urges predispose them to various sexual risky behaviors such as early sexual initiation, having sex with multiple partners, early marriage, unprotected sex, and premarital sexual practice [4].

Premarital sex is defined as voluntary sexual intercourse between unmarried persons with each other [5]. Globally, youths’ sexuality issue has become a recent concern in much research [5]. In developing countries, the prevalence of premarital sexual practice among youths is increasing tremendously from time to time [6].

Moreover, premarital sexual activity is a debatable issue in its normality. However, In most developing countries like Ethiopia, sexual activities done before a mirage are mostly done secretly because of fear of social disapproval [7,8]. On top of that in such kinds of communities, it is not socially acceptable for youths to buy and use contraceptives before they got married. Due to this, youths are invited to end up unprotected sex which in turn lets them be suffered from serious health problems such as unintended pregnancies, unsafe abortion, Sexually Transmitted Diseases (STDs), Human Immunodeficiency Virus (HIV) and even to death. [9–12].

The prevalence of premarital sexual practice is highly variable across regions in Ethiopia, which ranges from 22.2% to 71%. [11–17]. Even though there is an increase in youth’s sexual activity, preventive reproductive health behaviors, such as condom utilization, are not satisfactory among these group of population [12,14].

As different studies showed, premarital sexual practice is associated with various factors either positively or negatively. These factors include sex [14,17], older age and peer influence [16], media exposure and work to earn money [17], having a boy/girlfriend [12] residence and family [18], use of social media[14], comprehensive HIV knowledge, Alcohol and smoking [19].

Indeed, there are several studies done in different parts of the country concerning premarital sexual practice among youths. However, none of them were used nationally representative data. Therefore, this study aimed to assess the prevalence and determinants of premarital sexual behavior of Ethiopian youths based on the nationally representative data of 2016 Ethiopian Demographic and Health Survey (EDHS). Furthermore, determining the prevalence and identifying factors associated with premarital sexual practice would have great importance for policymakers at the national level to design effective and tailored sexual education programs.

## Methods and materials

### Study period and setting

A community-based crossectional study was conducted from 18, 2016, to June 27, 2016. The survey was conducted in all regions of Ethiopia. Ethiopia is the second-most populous country next to Nigeria in Africa with an estimated population of 114 million [20]. About 21% of the population is in the age group of 15-24 [21]. Regarding the health care system, Ethiopia has introduced a health sector transformation plan which is organized by a three-tier health-delivery service system lead by a ministry, the so-called ministry of health of Ethiopia. These metaphorical tires comprise the primary level (health posts, health centers, and primary hospitals), secondary level (general hospitals), and tertiary level (specialized hospitals) [22].

### Data source, Sampling technique, and population

This study was conducted based on the 2016 Ethiopian demographic and health survey data (EDHS). The survey used the Ethiopian Population and Housing Census, which was conducted in 2007 by the Ethiopia Central Statistical Agency, as a sampling frame. The frame was a complete list of 84,915 enumeration areas (EAs) in which each EAs covers an average of 181 households.

Moreover, EDHS followed two-stage stratified sampling technique. In the first stage, 645 EAs (202 in urban areas and 443 in rural areas) were selected whereas, In the second stage, 28 households per cluster were selected from the newly created household listing. All necessary information about the sampling strategy, questioner, or other important information is available in the 2016 EDHS report [23]. Finally, the total weighted samples of 7389 youths were included for this study’s analysis.

### Study variables

The outcome variable of this study was premarital sexual behavior, which is defined as ever had sexual intercourse before marriage. The independent variables for the present study were; participants age, sex, religion, residence, region, education, wealth index, internet use, mobile phone ownership, bank account ownership, substance use (smoking, alcohol drinking, khat, chewing), ever tested for HIV, and media exposure, all of which were extracted from the EDHS 2016 data.

### Data processing and analysis

Following data extraction, Data coding and transformations were done by using Stata version 14.2 software to make the data ready for analysis. Besides, sampling weight was done to adjust for non-proportional allocation of the sample to strata and regions during the survey process. Since the outcome variable was dichotomy (1=Yes and 0= No), binary logistic regression was fitted to identify important factors associated with premarital sex. Variables with p-value < 0.2 in the bivariable logistic regression analysis were entered into multivariable logistic regression analysis.

The Hosmer and Lemeshow test was done to assess the overall model fitness, it was 0.69 indicated overall good model fitness. Besides, multicollinearity was tested based on variance inflation factor (VIF) and we have got a VIF of less than five for each independent variable with a mean VIF of 1.98, indicated there was no significant multicollinearity between independent variables. Finally, p-value < 0.05, and 95% confidence interval were used to determine statistical significance.

## Ethical issue

Since we have used secondary data was not required. However, Permission to use the data was obtained from the DHS Program.

## Results

### Sociodemographic characteristics

The median (interquartile range) age of the participants was 18 (16-20) with the age range of 15 to 24. Majority (70%) of them were in the age group of 15-19. More than half (52.6%) of the participants were females. About 4409 (60%) and 749 (10.1%) of the participants had a primary education level and hadn’t attended any formal education at all respectively [Table 1].

**Table 1:** Sociodemographic characteristics of youths in Ethiopia (n = 7389)

| Variable | Frequency | Percent (%) |
| --- | --- | --- |
| Age |  |  |
| 15-19 | 5,169 | 70 |
| 19-24 | 2,220 | 30 |
| Sex |  |  |
| Male | 3, 500 | 47.4 |
| Female | 3,889 | 52.6 |
| Religion |  |  |
| Orthodox | 3336 | 45.1 |
| Muslim | 2012 | 27.2 |
| Protestant | 1897 | 25.7 |
| Other | 144 | 1.9 |
| Residence |  |  |
| Rural | 5482 | 74.2 |
| Urban | 1907 | 25.8 |
| Region |  |  |
| Agrarian | 6471 | 87.6 |
| Metropolitans | 584 | 7.9 |
| Pastorals | 334 | 4.5 |
| Education |  |  |
| No formal education | 749 | 10.1 |
| Primary | 4409 | 59.7 |
| Secondary | 1707 | 23.1 |
| Tertiary | 524 | 7.1 |
| Wealth index |  |  |
| Poor | 2007 | 27.2 |
| Medium | 1324 | 17.9 |
| Rich | 4058 | 54.9 |
| Owe bank account |  |  |
| No | 6353 | 86 |
| Yes | 1036 | 14 |
| Owe mobile phone |  |  |
| Yes | 3303 | 44.7 |
| No | 4086 | 55.3 |

### Media exposure and behavioral factors

From the total participants, only 943 (12.8%) of them had ever used the internet of which 358 (4.9%) of them used at least once a week. The majority of the participants (66.8%) exposed at least for one of the mass medias, where more than half (52%) of them were used to watch television. About 2733 (37%) and 801(11%) of the participants had ever drunk alcohol and chewed khat respectively. Moreover, only 2880 (39%) and 2489 (34%) of the participants know both methods (condom use and having only one sexual partner only) of HIV prevention and had ever tested for HIV respectively. Indeed, the prevalence of self-reported premarital sex was found to be about 11% (795) [Table 2].

**Table:** Summary statistics of factors of premarital sexual practice among Ethiopian youths (n = 7389)

| Variable | Frequency | Percent |
| --- | --- | --- |
| Internet use |  |  |
| Never | 6446 | 87.2 |
| Ever | 943 | 12.8 |
| Frequency of internet use |  |  |
| Not at all | 6446 | 87.2 |
| Less than once a week | 273 | 3.9 |
| At least once a week | 358 | 4.9 |
| Almost every day | 312 | 4.2 |
| Overall Media exposure |  |  |
| Exposed | 4937 | 66.8 |
| Unexposed | 2452 | 33.2 |
| Specific media exposure |  |  |
| Ever used to read newspaper/magazine | 2180 | 29.1 |
| Ever used to listen to the radio | 3580 | 48.5 |
| Ever used to watch television | 3832 | 51.9 |
| Ever alcohol use |  |  |
| No | 4656 | 63 |
| Yes | 2733 | 37 |
| Ever smoke cigarette |  |  |
| No | 7338 | 99.3 |
| Yes | 51 | 0.7 |
| Ever chewed Khat |  |  |
| No | 6588 | 89 |
| Yes | 801 | 11 |
| Heard about HIV |  |  |
| Yes | 6982 | 94.5 |
| No | 407 | 5.5 |
| Knowledge HIV on preventive methods |  |  |
| Know both methods | 2880 | 39 |
| Don't know | 4509 | 61 |
| Comprehensive HIV knowledge |  |  |
| No | 4163 | 56.3 |
| Yes | 3226 | 43.7 |
| Ever Heard of STI |  |  |
| Yes | 7009 | 94.8 |
| No | 380 | 5.2 |
| Ever tested for HIV |  |  |
| No | 4900 | 66.3 |
| Yes | 2489 | 33.7 |
| Awareness of planning methods |  |  |
| No | 254 | 3.4 |
| Yes | 7135 | 96.6 |

### Factors associated with premarital sexual practice

Independent variables with a p-value of less than 0.2 in the univariable logistic regression analysis were entered into multiple logistic regression analysis. These variables were, age, sex, education, residence, employment, region, mobile ownership, bank account ownership, wealth, internet, ever alcohol use, ever smoke, ever chew khat, and ever tested for HIV. Moreover, multivariable logistic regression analysis revealed that being in the age group of 15 – 24, male sex, being employed, being from the pastoral region, having a mobile phone, being an internet user, ever drink alcohol, ever chewed khat, and ever tested for HIV were positively and significantly associated with premarital sexual practice [Table 3].

**Table 1:** Factors associated with Premarital sexual practice among Ethiopian youths (n = 7389)

| Variable | Premarital sex |  | Bivariable analysis | Multivariable analysis |
| --- | --- | --- | --- | --- |
|  | Yes (n = 795) | No (n = 6594) |  |  |
|  | Frequency (%) | Frequency (%) | COR (95% CI) | AOR (95% CI) |
| Age |  |  |  |  |
| 15-19 | 272 (5.3) | 4897 (94.7) | 1 | 1 |
| 20-24 | 523 (23.6) | 1697 (76.4) | 5.6 (4.7,6.9) ** | 3.6 (2.8,4.6) ** |
| Sex |  |  |  |  |
| Male | 565 (14.5) | 3324 (85.5) | 2.4 (1.9, 3) ** | 1.7 (1.3,2.2) ** |
| Female | 230 (6.6) | 3270 (93.4) | 1 | 1 |
| Education |  |  |  |  |
| No education | 61 (8.2) | 688 (91.8) | 1 | 1 |
| Primary | 364 (8.3) | 4045 (91.7) | 1.01 (.67,1.5) | 1.01 (0.7,1.7) |
| Secondary | 218 (12.8) | 1489 (87.2) | 1.6 (1.1,2.5) * | 0.96 (0.6,1.6) |
| Tertiary | 252 (40.4) | 372 (59.6) | 4.5 (2.9,7.3) ** | 1.1 (0.6,1.9) |
| Residence |  |  |  |  |
| Urban | 312 (16.4) | 1595 (83.6) | 2 (1.6,2.5) ** | 1 |
| Rural | 483 (8.8) | 4999 (91.2) | 1 | 1.1 (0.8,1.6) |
| Occupation |  |  |  |  |
| Employed | 615 (13.3) | 4006 (86.7) | 2.2 (1.7,2.8) ** | 1.4 (1.03,1.8) * |
| Unemployed | 180 (6.5) | 2588 (93.5) | 1 | 1 |
| Region |  |  |  |  |
| Agrarian | 634 (9.8) | 5838 (90.2) | 1 | 1 |
| Pastorals | 35 (10.5) | 299 (89.5) | 1.1 (.9,1.3) | 1.8 (1.3,2.4) ** |
| Metropolitan | 126 (21.6) | 458 (78.4) | 2.5 (2.1,3) ** | 1.3 (0.9,1.7) |
| Religion |  |  |  |  |
| Orthodox | 446 (13.4) | 2890 (86.6) | 2.2 (.7,6.7) | 1 |
| Protestants | 167 (8.8) | 1730 (91.2) | 1.4 (0.4,4.3) | 1.5 (0.9,2.3) |
| Muslims | 172 (8.6) | 1839 (91.4) | 1.3 (0.4,4.1) | 0.98 (0.6,1.5) |
| Other | 10 (6.7) | 135 (93.3) | 1 | 0.64 (0.2,1.9) |
| Media exposure |  |  |  |  |
| Exposed | 256 (10.4) | 2196 (89.6) | 1.02 (0.83,1.3) |  |
| Not exposed | 539 (10.9) | 4398 (89.1) | 1 |  |
| Owe mobile phone |  |  |  |  |
| Yes | 570 (17.3) | 2733 (82.7) | 3.6 (2.9,4.5) ** | 1.7 (1.3,2.3) ** |
| No | 225 (5.5) | 3862 (94.5) | 1 | 1 |
| Has bank account |  |  |  |  |
| Yes | 241 (23.7) | 795 (76.3) | 3.1 (2.5,4) ** | 1.1 (0.8,1.5) |
| No | 554 (8.7) | 5799 (91.3) | 1 |  |
| Wealth index |  |  |  |  |
| Poor | 142 (7.1) | 1865 (92.9) | 1 | 1 |
| Medium | 98 (7.4) | 1226 (92.6) | 1.1 (.73,1.5) | 0.9 (0.6,1.4) |
| Rich | 555 (13.7) | 3503 (86.3) | 2.1 (1.6,2.7) ** | 1.4 (1,1.9) |
| Internet use |  |  |  |  |
| Never used | 534 (8.5) | 5787 (91.5) | 1 | 1 |
| Ever used | 261 (24.4) | 807 (75.6) | 3.5 (2.8,4.4) ** | 1.8 (1.3,2.5) * |
| Alcohol use |  |  |  |  |
| Yes | 457 (16.7) | 2277 (83.3) | 2.6 (2.1,3.2) ** | 2.4 (1.7,2.5) ** |
| No | 338 (7.3) | 4317 (92.7) | 1 | 1 |
| Cigarette smoking |  |  |  |  |
| Never smoke | 774 (10.5) | 6564 (89.5) | 1 | 1 |
| Ever smoke | 21 (41.2) | 30 (58.8) | 5.7 (2.3,11.4) ** | 1.6 (0.7,4) |
| Khat chewed |  |  |  |  |
| Never chewed | 638 (9.7) | 5950 (90.3) | 1 | 1 |
| Ever chewed | 157 (19.6) | 644 (80.4) | 2.7 (1.7,3) ** | 2.4 (1.6,3.5) ** |
| Knowledge of HIV preventive methods |  |  |  |  |
| Know | 314 (10.9) | 2566 (89.1) | 1.03 (0.8,1.3) |  |
| Don't know | 481 (10.7) | 4028 (89.3) | 1 |  |
| Comprehensive HIV knowledge |  |  |  |  |
| Don't had | 432 (10.4) | 3731 (89.6) | 1 |  |
| had | 363 (11.3) | 2863 (88.7) | 1.2 (0.9,1.4) |  |
| Ever tested for HIV |  |  |  |  |
| No | 487 (9.9) | 4413 (90.1) | 1 | 1 |
| Yes | 308 (12.4) | 2181 (87.6) | 1.3 (1.03,1.6) * | 1.3 (1.1,1.6) * |

\* p-value < 0.05, and \*\* p-value < 0.001

## Discussion

The present study aimed to assess the prevalence and determinants of premarital sexual practice among youths in Ethiopia. The overall magnitude of premarital sexual practice was 10.8% (95% CI, 10 – 11.5) indicating that for every 10 youth at least one of them had experienced premarital sexual practice. This study’s finding is lower than the studies done in Wollega, 28.4% [14], Tigray,19% [16], Debre Markos, 31.3% [12], and Yabello, 71.9% [11]. This discrepancy may be due to the reason that the present study is based on the national data that may reduce the prevalence because of the inclusion of communities with strong social disapproval of premarital sexual practice.

Concerning to the factors of premarital sexual practice, being in the age group of 24-29 years, male sex, employed, from the pastoral region, having a mobile phone, use of the internet, alcohol drinking, Khat chewing, and ever tested for HIV were positively and significantly associated predictor variables with premarital sexual practice.

In this study, participants with the age group of 20-24 years were 5.6 times more likely to be engaged in premarital sexual practice as compared to participants with the age group of 15 to 19 years. This finding is similar to a study from Wollega [16]. It may be due to that youth with older age may have independent decision-making ability and they may have their income to run things by themselves without family recognition and permission.

Being male sex was another predictor variable that was positively and significantly associated with having premarital sexual activity, where the odds of having premarital sexual intercourse was higher among this groups. This finding is in line with studies from different parts of Ethiopia [11,12,14,17,24]. This might be due to that females are expected to keep their virginity until marriage in most parts of the country. On top of that young males are free to talk about sexual activities with others than young females [25].

The premarital sexual practice was higher among employed participants by 40% as compared to unemployed participants. This is in line with studies from southern Ethiopia [12] and Bahir Dar [17]. This might be because working youth can generate enough money so that they can offer different invitations for the opposite sex and exploit them for sex. Also, youth who can generate their income can have the capacity to pay for sex.

The present study revealed that youths from the pastoral region were 1.8 times more likely to engage in premarital sexual practice than youths from agrarians. This is contradicted to the study from Eastern Ethiopia where youths from urban society were more likely to engage in premarital sexual practice [24]. This may be due to those people from the pastoral region are mobile as a result of this, youths from this region may have low awareness of the consequences of premarital sex even if it is unprotected.

Participants who had either chewed khat or used to drink alcohol were 2.4 times more likely to engage in premarital sexual intercourse than khat and/or alcohol non-users. This is similar to several studies done previously in the country [8,10,12–14]. This might be due to that youths may lose their decision-making ability after once they took substance like alcohol and khat, which ultimately lets them engage in premarital sexual practice. Usually, in such risky conditions, sexual practices are done without taking care of it like without using condoms and to have multiple sexual partners, which may then lead them to end up with unwanted/teenage pregnancy, and sexually transmitted infections [8].

Moreover, the probability of having premarital sexual intercourse was 70% and 80% higher among participants who have a mobile phone and exposed to the internet respectively. This might be due to those participants having a mobile phone and internet access are obviously at higher risk to be exposed to sexually explicit materials, which in turn may predispose them to have premarital sex [17]. Hence, it is important to make internet services and mobile uses to be more selective and to use such services. Indeed, awareness of the consequences of inappropriate use of the internet and mobile phone has to be created.

In the present study, the odds of premarital sex were 1.3 times higher among youths who had ever tested for HIV, indicating that those who had tested for HIV were more likely to engage in premarital sexual activity than their counterparts. This is in line with a study from Wollega [16]. The possible reason might be due to a common belief that if youths know their HIV status each other, they are more likely to engage in sexual practice such as unprotected sex.

## Limitation of the study

One of the limitations of the present study is its crossectional nature that precludes us to conclude the causality of the observed associations. In addition to this, recall bias and social desirability bias may affect our estimation since the data was based on self-reports and the issue is socially sensitive. However, with the aforementioned limitations, the present study tried to picture out the national prevalence of the premarital sexual practice and its determinants among youth population of the country, which can be consumed by policymakers at the national level concerning youth’s reproductive health promotion and disease prevention.

## Strength of the study

In the presence of the aforesaid limitations, the present study addressed the determinants and prevalence of premarital sex based on nationally representative data so that policymaker or program implementers at the national level can use the evidence generated by the study for the promotion of reproductive health of youths in the country.

## Conclusion

For every 10 youths, at least one of them engaged in premarital sexual intercourse, which is non-negligible. Being in the age group of 20-24, male sex, employed, from the pastoral region, having a mobile phone, ever use of the internet, alcohol drinking, khat chewing, and ever tested for HIV were significant factors associated with the premarital sexual practice.

## Recommendation

National sexual education and reproductive health behavior change programs would have paramount importance in reducing the occurrence of premarital sex and ultimately, its consequences. Moreover, such intervention should give due emphasis to youths who use substance, have internet access, owe mobile phone and who are from pastoral regions. Indeed, adequate sexual counseling should be given about premarital sexual intercourse when youths the come for HIV test.

## Data Availability

Data used for the analysis is avialiable at DHS measure.

https://www.dhsprogram.com

## Acknowledgments

We would like to thank the MEASURE DHS for providing the data set for the study.

## Abbreviations

AOR: Adjusted Odds Ratio
AIDS: Acquired Immuno-Deficiency Syndrome
CI: Confidence Interval
COR: Crude Odds Ratio
EAs: Enumeration Areas
EDHS: Ethiopian Demographic, and Health Survey
HIV: Human Immune Virus
STDs: Sexually Transmitted Diseases

## Author’s Contribution

KSG conceived and designed the study, conducted processing and analysis, discuss the data, and drafted the manuscript for publication. AK and MW analyze and interpret the data, and reviewed the manuscript for its intellectual contents. All authors read and approved the final manuscript.

